# Reduced maternal healthcare interactions with general practice in the postnatal period during the COVID-19 pandemic, a cohort study of Greater Manchester residents

**DOI:** 10.64898/2026.08.18.26360757

**Authors:** Chantelle Cornett, George Tilston, Glen Martin, Victoria Palin

**Author notes:** Corresponding author: Victoria Palin. These authors contributed equally to this work.

## Abstract

**Background:** Maternal postpartum checks with a general practitioner (GP) are recognised as an essential service in England and vital for recovery after pregnancy and reducing risk of long-term morbidity. Despite this, its reported fewer than of women have a record of the examination in the recommended 6-8 weeks, with observed disparities in uptake nationally. The impact of the COVID-19 pandemic disrupted delivery of these checks nationally, but there is limited data on the impact of the pandemic and its recovery for regional populations representing diversity and areas of dense poverty and ethnic minority populations. This study utilised region level data to assess the impact of COVID- 19 on postnatal care.

**Methods:** Anonymised electronic health records with clinical coded birth events for females, aged 16-49 years, were analysed for patients registered with a GP using the Greater Manchester Care Record (GMCR) between January 2018 and August 2023. Unique delivery episodes were defined and monthly rates calculated separately for women with a postnatal-related code within 4-, 6-, 8-, or 12- weeks or 1 year follow-up. Rates were also generated by key maternal demographics to assess any differences in postpartum care. Interrupted time series, modelling the onset of the pandemic estimated the IRR of 0.49 (95% CI 0.40–0.58). To assess the impact of maternal characteristics on the odds of non-attendance at examination, a logistic regression adjusting for various maternal characteristics was fitted.

**Results:** There were 114,874 unique delivery episodes, relating to 85,076 women in the 12-week follow up cohort; 72,595 episodes to 55,784 women in 8-weeks and 28,846 episodes to 24,018 women in 6-weeks. The rate of postpartum checks was greater the longer the follow-up period. For checks within 8 weeks the first lockdown reduced from ∼325 per 1000 delivery episodes in 2019 to 225 per 1000 by April 2020 (30.8%), which remained low, before returning to pre-pandemic rates by rates by October 2022. Rates remained lower overall for Black, or Asian women compared to White.

**Conclusion:** The COVID-19 pandemic reduced postnatal follow-up in primary care across Greater Manchester, with rates frequently falling outside the recommended 6–8 week window. Significant disparities exist in the provision and uptake of these services. Improved integration of data across care sites, combined with enhanced risk management, could increase equity in access and support the timely delivery of care for those at greatest risk of postnatal complications and longer-term health issues.

## Introduction

The majority of pregnancy-related deaths during, or up to six weeks postpartum, are attributed to medical and mental health conditions exacerbated by pregnancy, with less than one-third directly related to obstetric complications [1, 2]. This highlights the importance of optimising women’s health before, during, and after pregnancy [2], and the importance of the maternal examinations in the early postpartum period with a GP. In England, UK, the Six to Eight Week check (SWC) should address recovery from birth, review both physical and mental health, pregnancy-related conditions requiring ongoing care, pelvic health, and family planning [3, 4]. As articulated in the Women’s Health Strategy for England (2022), and in the recent renewal (2026), timely and high-quality postnatal care is central to a life-course approach to women’s health, with the potential to prevent the onset of pregnancy-related morbidity in subsequent years [5, 6]. This is particularly pertinent for women with pre-existing or pregnancy-induced cardiometabolic conditions such as hypertension and diabetes, for whom a timely and thorough SWC is critical in reducing their long-term risk of chronic morbidity and mortality.

Despite its importance, evidence suggests the SWC is frequently delayed or missed entirely. A review covering 2015–2018 (pre-COVID-19 pandemic) found only 62% of women had a documented record of a SWC within 12 weeks of delivery, and just 40% within the recommended 6–8 weeks [7], with younger mothers, those with preterm deliveries, and women in more deprived areas of England were most likely to experience delayed or no check. In February 2020, an amendment to the GP contract formalised the SWC as an essential service in England, with £12 million in additional funding allocated to support its delivery [8]. However, the simultaneous onset of the COVID-19 pandemic significantly disrupted primary care services [9], likely stalling the intended impact of this policy change. At a national level, Hayes et al. (2025) demonstrated using OpenSAFELY data from over 2,500 practices that rates of postnatal examinations within 8 weeks fell by 24.1% between January and March 2020, recovering to above pre-pandemic levels by early 2023 [10]. Despite this recovery, significant inequalities persisted, with women in the most deprived quintile 34% more likely to miss a timely check compared to the least deprived, and elevated odds also observed among women of Asian or Asian British ethnicity and those in the North East and West Midlands [10].

Greater Manchester, England, represents an important setting in which to examine these dynamics further. With 28.7% of residents from ethnic minority groups, exceeding the national average of 26.5% [11], and 34.2% of the population falling within the lowest socioeconomic classification compared to 30.6% nationally [12], the region’s diversity and deprivation profile suggest it may be disproportionately affected by the inequalities identified at a national level. This study utilised the Greater Manchester Care (GM) Record (GMCR)[13], a region-wide integrated digital care system covering approximately 3.2 million residents and accessed by over 500 health and social care organisations, incorporating primary care interactions, hospital stays, and death records. The completeness and breadth of the GMCR provides a unique opportunity to examine postnatal care utilisation across a diverse regional population. The current study aimed to evaluate the overall rate of postnatal examinations within the recommended six to eight week window, stratified by deprivation, ethnicity, and region, and to investigate associations between women’s characteristics and the risk of a delayed or missed check before, during, and after the COVID-19 pandemic.

## Methods

### Data Source

The Greater Manchester Care Record (GMCR) was used to access routinely collected primary care electronic health records between 1^st^ March 2012 and 1^st^ August 2023. De-identified records were extracted for all females, aged 14-49 years, registered with a GM general practice (GP) with 2 or more maternity-related clinical codes on their records.

### Code lists

Primary care services within the GMCR are primarily compliant with the NHS standard of SNOMED- CT clinical terminology. Maternity-related clinical codes, as validated in published studies and used to identify pregnancy episodes within UK primary electronic health records [14], were converted to SNOMED CT using the NHS Technology Reference Update Distribution website [15] before data curation (22 June 2022)]. Further details on data curation are available in the GitHub Repository [16].

### Code check

All maternity-related clinical events relating to the concept of deliveries or postnatal examinations were tabulated separately, reviewed and refined by the study team so that only definitive delivery and postnatal codes remained in the analyses after the conversion. For example, “unspecified hypertension complicating pregnancy |childbirth and the puerperium unspecified” was removed from codes that indicated delivery events because it cannot reliably distinguish between the antepartum (before birth), intrapartum (during birth), or the puerperium (up to 8 weeks postnatally). Following review, the frequency of delivery and postnatal related codes were tabulated for the cohort (see, Supplementary Table 1).

### Defining discrete deliveries

The starting cohort included all female patients with two or more antenatal related coded events within the study period (n = 279,295). To identify unique pregnancies that likely resulted in a birth ≥ 22 weeks, the cohort was restricted to patients with at least one delivery code on their primary care record (n = 176,041). For each patient, delivery codes were ordered by date and the interval between consecutive delivery dates calculated. As ovulation can return approximately four weeks postpartum, the earliest biologically possible interval between one birth and a subsequent viable birth (>22 weeks); delivery codes separated by > 182 days were classified as distinct birth events.

Overall, 50.8% of patients had a single recorded delivery code. Among patients with multiple delivery-related codes, short-interval events were examined to identify implausibly proximate deliveries: 27.5% of patients had delivery-code intervals within 182 days, of which 75% occurred within 49 days, and median interval of 14 days. These were therefor interpreted as coding of the same delivery over distinct delivery episodes. Across the 176,041 patients with at least one delivery, there were 251,770 distinct episodes with a mean of 1.43 deliveries per patient (IQR 1-2, max 9).

From the earliest unique delivery episode, patient records were then examined for recordings of postnatal codes within predefined follow-up periods. Five follow-up windows were assessed: up to 4, 6, 8, or 12 weeks, and up to a maximum of 1 year postpartum.

### Outcome and exposures

The primary outcome was the rate of postnatal health care interactions within 4-, 6-, 8-, or 12-weeks postpartum to a maximum of one year following a coded birth events in primary care. Secondary outcomes included any disparities in postnatal health care interactions based on ethnicity, socioeconomic status, or regional variations within Greater Manchester. The exposure of interest was the impact of the COVID-19 pandemic period postnatal examinations, defined as the period encompassing the substantive phase of COVID-19 pandemic restrictions in England, from 1st January 2020 to 31st December 2021 [17] with a pandemic recovery defined from 1^st^ January 2022 to the end of follow-up.

### Data Processing

#### Statistical analyses

The most frequent delivery and postnatal examination codes were first tabulated. Similarly, the cohorts characterised were described, stratified across each follow-up window. Monthly rates were calculated overall and by predefined subgroups including maternal ethnicity, index of multiple deprivation (IMD) quintile and region, for each window and visualised as time-series plots.

Negative binomial interrupted time-series (ITS) was used to examine the impact of the COVID-19 pandemic on the rate of postnatal examinations. Monthly counts for postnatal examinations were modelled, with an offset for the population size of those with a birth-related code up to 8 weeks before, adjusting for a binary variable to indicate the onset of COVID-19 (January 2020), a continuous monthly count variable, and time since the interruption variable. The ITS focused specifically on the maximum recommended window for postnatal examinations (within 8 weeks postnatally) to estimate the incidence rate ratio (IRR) before and after the onset of the first national lockdown. The counterfactual was estimated following the start of the pandemic modelling the rates of postnatal examinations if there was no interruption by the COVID-19 pandemic.

Logistic regression analyses were used to estimate the impact of maternal characteristics including age, ethnicity, deprivation measured with an Index of Multiple Deprivation (IMD) quintile, and region of residence on the risk of no postnatal examination. BMI was not included due to high volume of missingness and given as little as 2% ethnicity was not reported, a complete case approach was adopted. Interaction terms between the COVID 19 period and key covariates were also tested to assess whether the effect of the pandemic differed across demographic subgroups; Where interaction terms are included, main effect coefficients represent the effect of COVID-19 period for the pre-pandemic period only and are not directly comparable to estimates from the main effects model.

## Results

There was a total of 251,770 distinct delivery events to 176,041 patients (mean 1.43). 388 delivery codes were present within the cohort, following review 338 unique codes were included in the analysis, the most common being “spontaneous vaginal delivery”, “spontaneous vertex delivery”, and “svd – spontaneous vaginal delivery”. 306 postnatal codes were present within the cohort, following review 95 unique codes were included in the analysis, the most common being “puerperal depression”, “postpartum care” and postnatal maternal examination” (Supplementary Table 1) Of these codes, while included 0.78% of codes related to the eight-week baby check appointment, 0.91% referred to postnatal care that might occur elsewhere in the community, and 14.1% were related to breastfeeding information recorded as part of the postnatal examination, the baby check appointment, or a combination.

The proportion of deliveries with a postnatal examination increased with longer follow-up window. Overall, the percentage of deliveries with a postnatal coded examination up to one year postpartum was 48.4%. For those with postnatal examinations, the mean number of days from delivery was 54.4 days (IQR 43-61). Just 5.4% of postnatal examinations occurred within 4 weeks of a birth, 11.5% to 28.8% within the recommended 6-8 weeks and 45.6 to 48.6% within 12 weeks and 12 months, respectively (Table 1).

**Table 1.** Overall number of deliveries and number with postnatal examinations within the follow-up period.

|  |  |  |
| --- | --- | --- |
|  | **N** | **%** |
| **Total Deliveries** | 251,770 |  |
| **Postnatal Examination follow-up period** |  |  |
| 4 weeks | 13,702 | 5.4 |
| 6 weeks | 28,846 | 11.5 |
| 8 weeks | 72,595 | 28.8 |
| 12 weeks | 114,874 | 45.6 |
| 12 months | 121,940 | 48.4 |

Prior to the pandemic, 8-week postnatal examination rates had already been declining, from 34.8% in 2013 to 27.9% in 2019. The onset of COVID-19 accelerated this decline, with rates falling to 22.2% in 2020. Although some recovery was observed by 2022 (23.4%), rates had not returned to expected pre-pandemic levels by the end of the study period.

A negative binomial interrupted time-series analysis showed that the rate of postnatal examinations within 8 weeks of a birth-related code was lower following the onset of the COVID-19 pandemic than would have been expected based on pre-pandemic trends (Figure 1). Prior to January 2020, the rate of postnatal examinations decreased over time (IRR slope: 0.997, 95% CI: 0.9965, 0.9974). At the point of interruption, there was an immediate decrease in the rate of postnatal examinations (IRR 0.49, 95% CI: 0.40-0.58). In the post-interruption period, the monthly trend increased relative to the pre-pandemic trend (IRR slope 1.01, 95% CI: 1.00-1.01). Overall, observed rates of postnatal examinations after the onset of the pandemic were lower than the counterfactual estimates, suggesting that the COVID-19 pandemic was associated with a sustained change in postnatal examination rates over the observation period.

**Figure 1.**
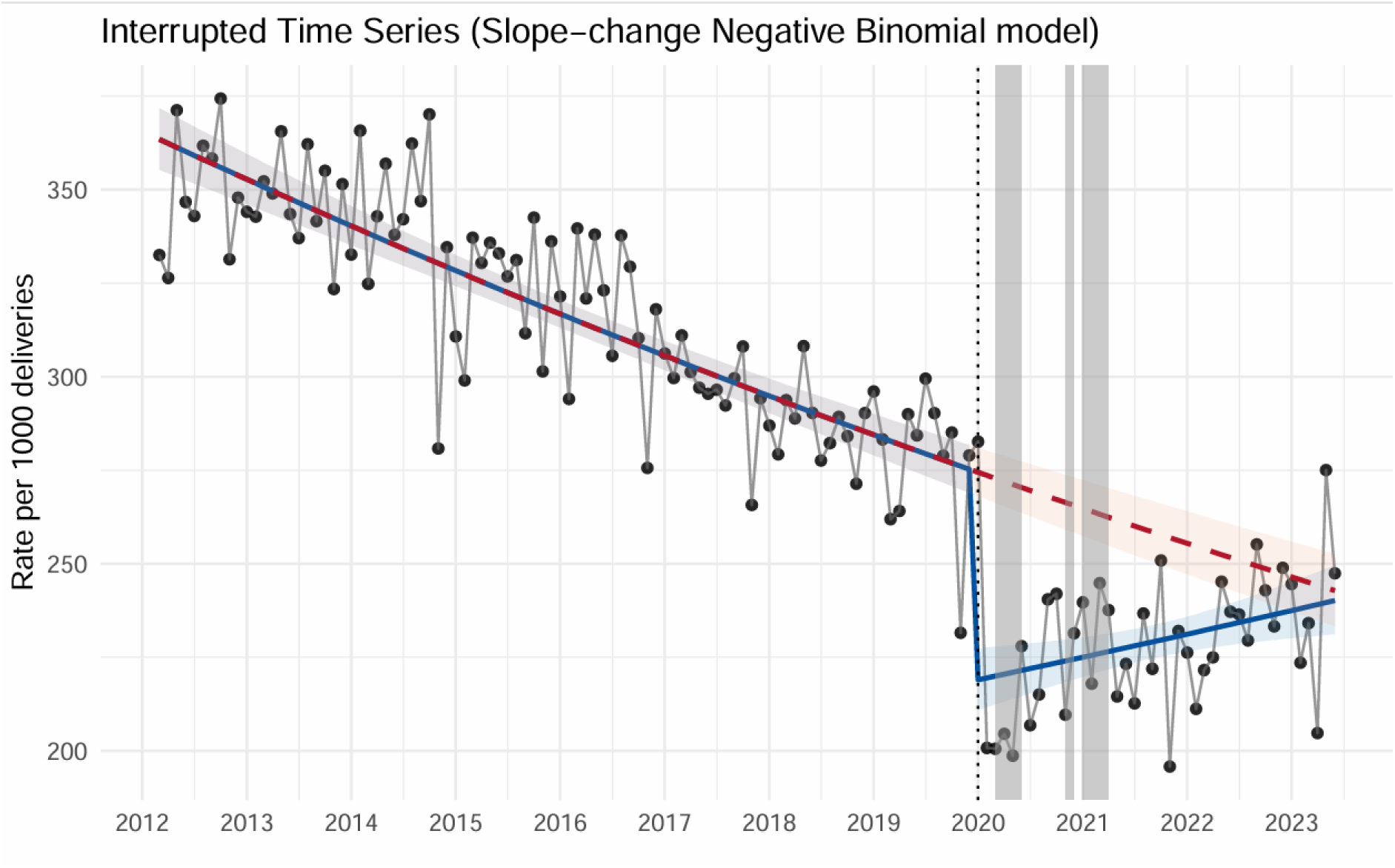
Negative Binomial interrupted time series model.

Across all windows the mean age was 30 years. 16.7% of individuals identify as Asian or Asian British, 5.0% as Black or Black British, 2.3% as Mixed race, 12.0% of other ethnic origin and 61.9% as White. Individuals lived in areas with an IMD quintile of (most-least deprived) at rates of 46.7%, 18.6%, 11.6%, 12.6% and 10.5% respectively. The cohort characteristics for each follow-up window are summarised in Table 2. Baseline characteristics (Table 2) and logistic regression analyses (Table 3) revealed differences between those who attended postnatal checks and those who did not. Age was generally similar between those who attended and those who did not across all time points, though logistic regression confirmed a small but statistically significant association, with increasing age slightly reducing the odds of missing a postnatal check (OR 0.993, 95% CI 0.991–0.994, p<0.001). BMI was not included in the model due to high levels for missingness.

**Table 2.** Number of deliveries with postnatal examinations (and % of overall deliveries) within the follow-up period by ethnic group, index of multiple deprivation, practice region and smoking status.

|  | 6 weeks |  | 8 weeks |  | 12 weeks |  |
| --- | --- | --- | --- | --- | --- | --- |
|  | N | % | N | % | N | % |
| **Ethnic Group:** |  |  |  |  |  |  |
| Asian or Asian British | 4273 | 10.1 | 10221 | 24.3 | 17327 | 41.1 |
| Black or Black British | 1416 | 11.2 | 3079 | 24.3 | 5053 | 40.0 |
| Mixed | 655 | 11.3 | 1456 | 25.2 | 2329 | 40.3 |
| Other | 3409 | 11.3 | 8488 | 28.1 | 13522 | 44.8 |
| White | 18518 | 11.9 | 47821 | 30.7 | 74140 | 47.6 |
| Refused/Not Stated | 570 | 10.8 | 1511 | 28.6 | 2471 | 46.7 |
| **IMD:** |  |  |  |  |  |  |
| 1 | 11657 | 9.9 | 28217 | 24.1 | 47663 | 40.6 |
| 2 | 5610 | 12.0 | 13787 | 29.5 | 21834 | 46.7 |
| 3 | 3627 | 12.5 | 9356 | 32.1 | 14606 | 50.2 |
| 4 | 4373 | 13.8 | 11202 | 35.5 | 16282 | 51.5 |
| 5 | 3511 | 13.3 | 9891 | 37.3 | 14261 | 53.8 |
| **Practice Region:** |  |  |  |  |  |  |
| Bolton | 3233 | 12.0 | 8568 | 31.8 | 13652 | 50.7 |
| Bury | 1396 | 9.1 | 4401 | 28.7 | 8538 | 55.8 |
| HMR | 1910 | 9.4 | 5003 | 24.5 | 7969 | 39.1 |
| Manchester | 6609 | 13.1 | 12757 | 25.3 | 7969 | 39.1 |
| Oldham | 2204 | 10.2 | 5358 | 24.8 | 8743 | 40.4 |
| Salford | 2678 | 12.8 | 6270 | 29.9 | 9233 | 44.0 |
| Stockport | 2820 | 10.8 | 7377 | 28.2 | 12641 | 48.3 |
| Tameside Glossop | 1276 | 7.8 | 2770 | 17.0 | 6731 | 41.3 |
| Trafford | 2456 | 12.1 | 7077 | 34.8 | 9840 | 48.3 |
| Wigan | 3492 | 13.0 | 11159 | 41.6 | 14986 | 55.9 |
| **Worst Smoking Status:** |  |  |  |  |  |  |
| Non-Smoker | 16433 | 11.6 | 41387 | 29.2 | 65544 | 46.3 |
| Non-Trivial Smoker | 12207 | 11.4 | 30770 | 28.6 | 48486 | 45.1 |
| Trivial Smoker | 69 | 11.4 | 157 | 26.0 | 259 | 42.9 |
| Unknown | 137 | 6.5 | 281 | 13.4 | 585 | 27.8 |

**Table 3.**
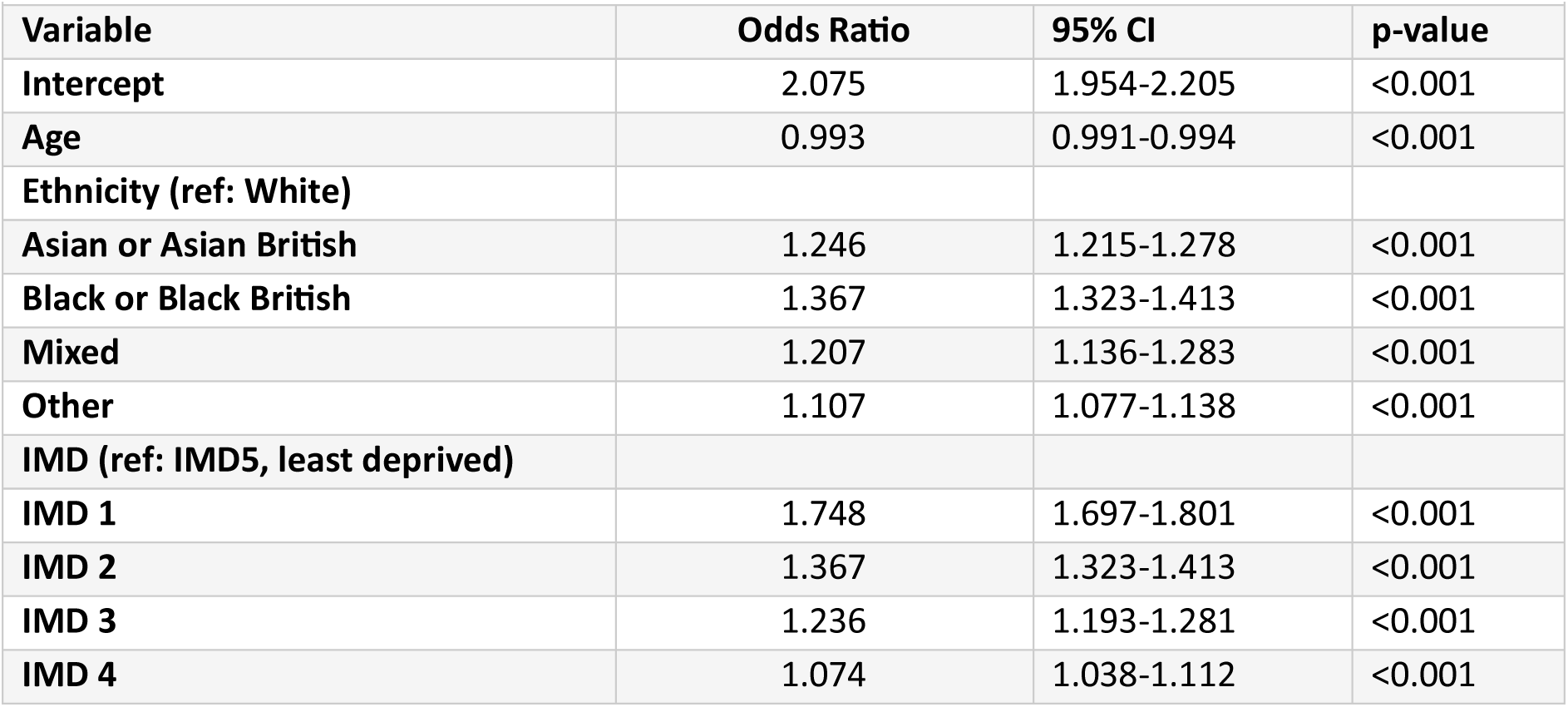
Logistic Regression Model: Odds of No Postnatal Examination within 8 Weeks.

The COVID-19 pandemic significantly affected postnatal check rates. Attendance dropped markedly in April 2020 (Figure 2), particularly in deprived and minority ethnic groups (Figures 3-4). Although rates began to recover by 2022, the pandemic’s impact on postnatal care was more pronounced among vulnerable groups, with recovery varying by IMD and ethnicity (Figures 3-4). Logistic regression (Table 3) confirmed that women delivering during the acute pandemic period (2020– 2021) had significantly higher odds of missing a postnatal check compared to the pre-pandemic period (OR 1.063, 95% CI 1.040–1.086, p<0.001). By the recovery period (2022 onwards), odds of missing a check were slightly reduced compared to pre-pandemic levels (OR 0.959, 95% CI 0.936– 0.983, p<0.001), suggesting partial but incomplete recovery in postnatal care attendance.

**Figure 2.**
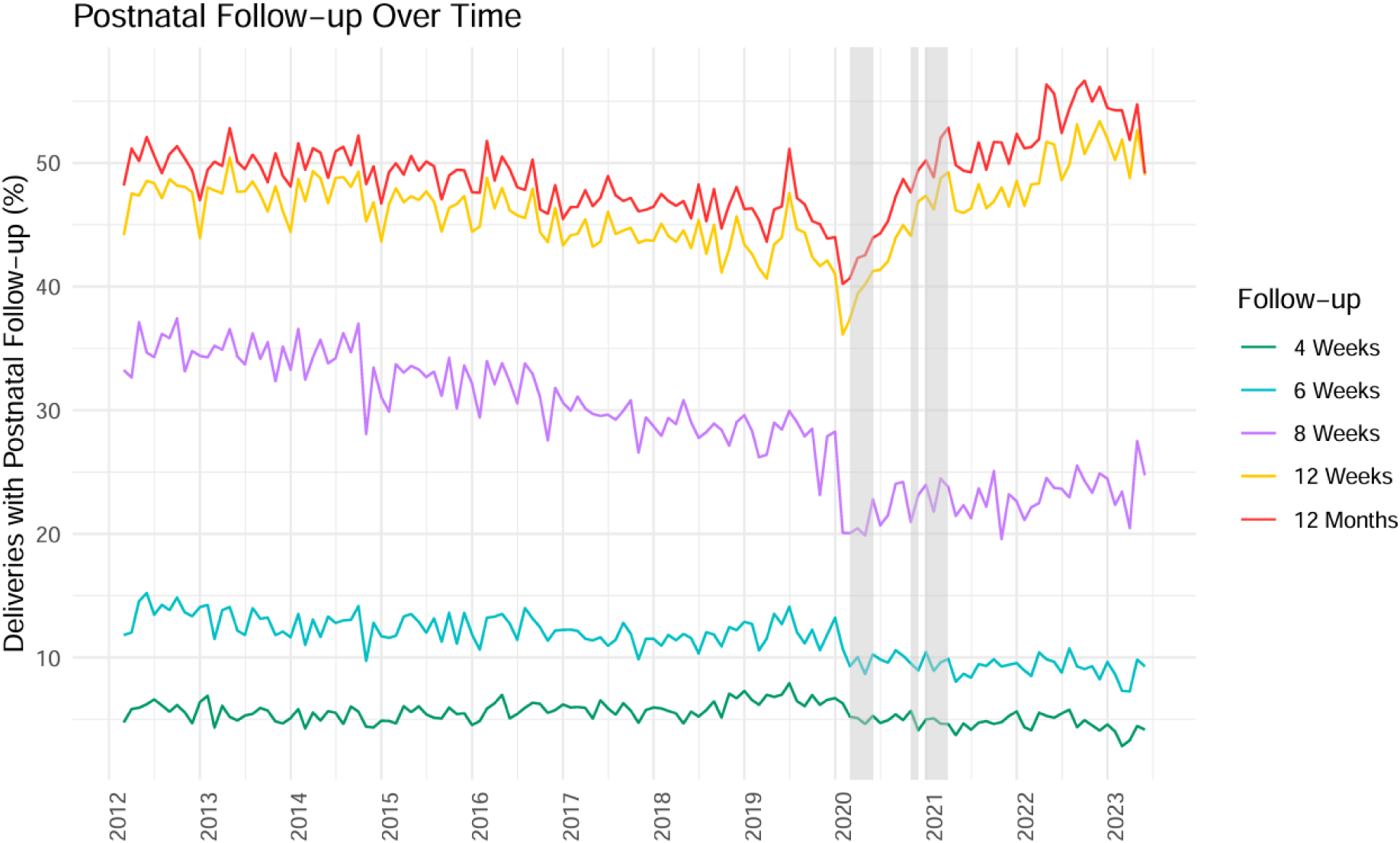
Deliveries with postnatal follow-up over time from 2012 to 2023.

**Figure 3.**
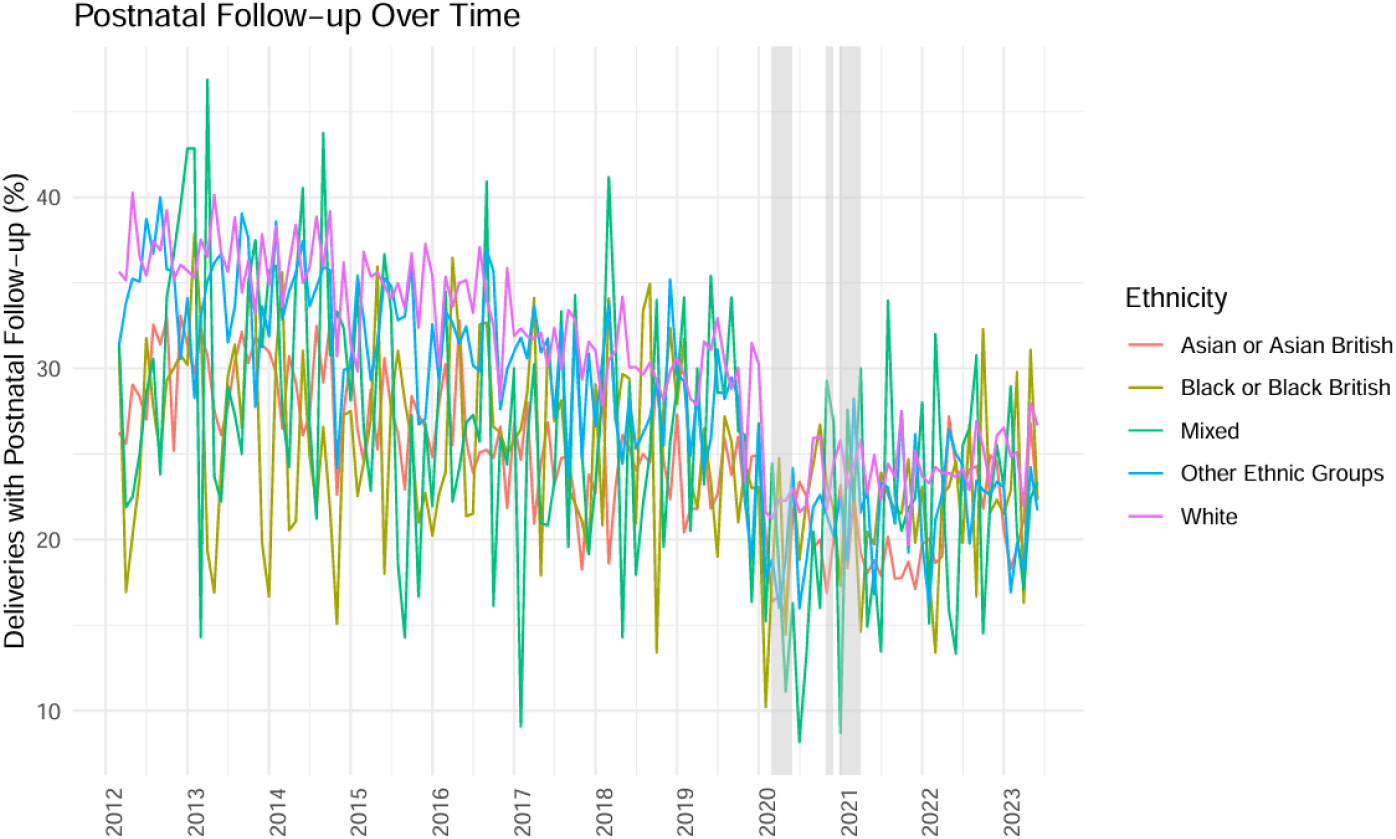
Deliveries with postnatal follow-up over time by ethnicity from 2012 to 2023.

**Figure 4.**
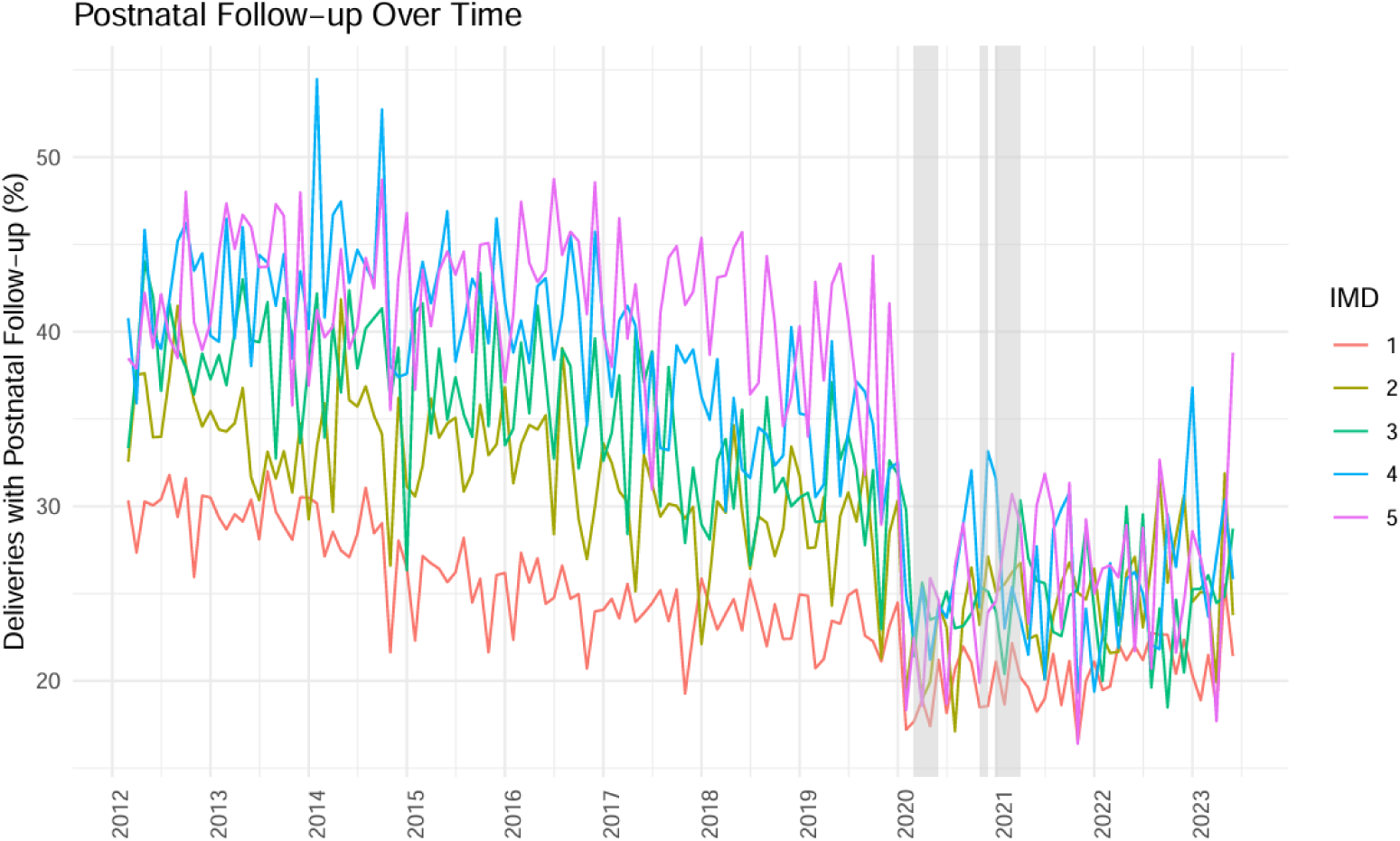
Deliveries with postnatal follow-up over time by IMD quintile from 2012 to 2023.

Ethnic disparities in postnatal attendance were evident across all time points (Figure 3). White women had the highest attendance rates at 8 weeks (30.7%), compared to Asian or Asian British (24.3%), Black or Black British (24.3%), Mixed (25.2%), and other ethnic groups (28.1%), with these disparities persisting across the 6 and 12 week cohorts (Table 2). Logistic regression (Table 3) confirmed that for the 8-week cohort, all minority ethnic groups had significantly higher odds of missing a postnatal check compared to White women, with Asian or Asian British ethnicity women having the greatest odds (OR 1.246, 95% CI 1.215–1.278, p<0.001), followed by Mixed (OR 1.207, 95% CI 1.163–1.283, p<0.001), Black or Black British (OR 1.168, 95% CI 1.119–1.220, p<0.001), and other ethnic groups (OR 1.107, 95% CI 1.077–1.138, p<0.001).

Significant regional variations in postnatal check attendance were observed within Greater Manchester. Wigan had the highest attendance (41.6% in the 8 week cohort), while the Tameside Glossop practice region had the lowest rate (17.0%), highlighting regional disparities in care access (Table 2, Supplementary Figure 1).

Socioeconomic disparities were also evident (Table 2, Figure 4), with women from more deprived areas less likely to attend checks. Women in the least deprived quintile (IMD 5) had the highest attendance at the recommended 8 weeks (37.3%), compared to 24.1% among those in the most deprived quintile (IMD 1). Logistic regression (Table 3) reinforced this pattern, with women in the most deprived quintile (IMD 1) having substantially higher odds of missing a postnatal check compared to those in the least deprived quintile (OR 1.748, 95% CI 1.697–1.801, p<0.001). A clear deprivation gradient was observed, with odds of non-attendance decreasing progressively across IMD quintiles (IMD 2: OR 1.367, 95% CI 1.323–1.413; IMD 3: OR 1.236, 95% CI 1.193–1.281; IMD 4: OR 1.074, 95% CI 1.038–1.112), confirming that socioeconomic deprivation is a significant independent predictor of non-attendance at postnatal checks.

A further logistic regression model was fitted with interaction terms between COVID-19 period and ethnicity as well as deprivation to assess whether the pandemic’s impact differed across subgroups (Supplementary Table 2). It showed a significant underlying increase in non-attendance over time (OR 1.054, 95% CI 1.049–1.059, p<0.001). During the acute pandemic period, no significant ethnicity-by-COVID interactions were observed, suggesting the pandemic affected all ethnic groups similarly. However, during the recovery period, Asian or Asian British and Black or Black British women showed significantly greater recovery in attendance relative to White women (OR 0.846, 95% CI 0.783–0.913, p<0.001 and OR 0.872, 95% CI 0.771–0.988, p=0.030, respectively).

For deprivation (Supplementary Table 2), a consistent pattern emerged whereby the pandemic’s impact on non-attendance was attenuated among more deprived women. This was significant for IMD 1–3 during the acute period (IMD 1: OR 0.700, p<0.001; IMD 2: OR 0.739, p<0.001; IMD 3: OR 0.811, p<0.001) and for IMD 1–2 during recovery (IMD 1: OR 0.693, p<0.001; IMD 2: OR 0.731, p<0.001).

## Discussion

This study provides valuable insights into postnatal care utilisation in Greater Manchester, highlighting significant disparities by socioeconomic status, ethnicity, and region, especially in the context of the COVID-19 pandemic. Overall, just 48.4% of deliveries had a recorded postnatal examination within one year, with only 28.8% occurring within the recommended 8-week window. Our findings illustrate a pronounced reduction in postnatal check attendance following the onset of the pandemic, with an immediate 51% reduction in the rate of postnatal examinations at the point of interruption (IRR 0.49, 95% CI 0.40–0.58), particularly among women from deprived areas and minority ethnic groups. Although attendance rates showed signs of recovery over time, these disparities remained persistent, emphasising the need for targeted interventions to improve postnatal care access and attendance among vulnerable populations.

The COVID-19 pandemic severely disrupted postnatal care delivery in Greater Manchester, aligning with previous reports of disruptions to maternity care during the pandemic [10], likely due to healthcare service reallocation, restricted in-person visits, and altered health-seeking behaviours. Notably, the immediate decline observed in Greater Manchester was considerably steeper than that reported nationally by Hayes et al. (2025), which may reflect the region’s particularly high burden of COVID-19 during the first wave, compounded by its concentration of socioeconomic deprivation and ethnic minorities [18], which can hinder access to postnatal checks. Additionally, systemic barriers, including limited culturally appropriate services, may further disadvantage ethnic minority groups. Addressing these social determinants of health is crucial to promoting equitable access to postnatal care. Crucially, our data also suggest the pandemic accelerated a pre-existing downward trend rather than an isolated disruption. Despite partial recovery, rates remained substantially below pre-pandemic levels throughout the observation period.

Ethnic disparities in postnatal attendance were evident across all follow-up windows. Systemic factors such as differential access, cultural barriers, and potential discrimination within healthcare settings may contribute to these persistent disparities. Previous literature has shown a lack of awareness surrounding the existence of the postnatal check, both among healthcare professionals and women [19], and culturally sensitive outreach and education programmes remain essential to improving postnatal care uptake among minority ethnic women.

Socioeconomic disparities were also significant, with a clear deprivation gradient observed across IMD quintiles. Importantly, interaction analyses (Supplementary Table 2) revealed that the additional impact of the pandemic on non-attendance was attenuated among more deprived women, particularly during the acute period, likely reflecting a ceiling effect whereby attendance was already markedly lower in these groups prior to the pandemic. This pattern persisted into the recovery period for the two most deprived quintiles, suggesting that post-pandemic recovery in postnatal care has been less equitable across socioeconomic groups.

Regional disparities were apparent, with variations likely reflect differences in local healthcare capacity, population density, and maternal health resources. Future research exploring regional service provision and patient satisfaction with postnatal care could inform strategies to address these regional disparities. Enhancing the integration of services and standardising care practices may help reduce regional inequalities in postnatal care. Looking at other maternity services, it has been shown that there are regional and ethnic disparities in the uptake of a baby postnatal check [20]. Whilst the objective of this paper is focusing on maternal postnatal checks, it is more likely that those with no baby postnatal check are also not receiving maternal postnatal checks.

The implications of this study for practice and policy are significant. Targeted interventions are necessary to address socioeconomic and ethnic disparities in postnatal care access. Community- based outreach, mobile health units, and telehealth services could improve access for women facing barriers to in-person care. Broader public health approaches addressing social determinants of health such as housing, education, and income support, are essential to reducing these inequities.

Policymakers should prioritise investments in services that enhance postnatal care access for underserved populations, ensuring care is equitable and culturally appropriate.

Integrating postnatal care within broader maternal health services is critical, not only to ensure continuity of care during public health emergencies but also to support preventative medicine. The timely conduct of postnatal checks provides an opportunity to identify and manage potential pregnancy-related health complications before they develop into longer-term conditions. This can include addressing hypertension, diabetes, or mental health concerns, alongside promoting lifestyle modifications such as improved nutrition and physical activity. Establishing robust protocols for maintaining essential maternal services during crises can prevent the significant disruptions observed during the pandemic. Furthermore, enhanced data-sharing across healthcare providers could facilitate better monitoring of care utilisation and outcomes, enabling timely interventions for at-risk populations and contributing to the overall health of mothers and their families.

The study’s strengths include the use of large, region-specific data and comprehensive insights into postnatal care patterns across Greater Manchester. However, limitations include the observational design, which restricts causal inferences, and potential confounding factors, such as maternal health status, that were not accounted for. While the study adjusted for key variables, residual confounding may still influence the findings. For example, the inclusion of alive patients only may have resulted in a relatively ’healthier’ population. However, as the target population consisted of younger women of childbearing age (and <1% of childbearing women within the data died), this is unlikely to have introduced significant bias. In addition, a short follow-up means our results are less likely to be impacted by loss to follow-up. Although more records are missing from diverse populations, the amount of missing data was limited (<2% for ethnicity unknown and <1% for IMD unknown). In addition, there may be bias towards the null due to underestimation of the true effects of ethnicity and deprivation on the rate of postnatal checks. This would be caused by GPs not coding events correctly. The incorrect coding of events may also lead to the misidentification of pregnancy events. To determine the effect of incorrect coding, we would need to investigate other factors such as comorbidities and complications in pregnancy, but due to the structure of primary care data it is currently too difficult to determine exact pregnancy start and end dates to be able to do so.

## Conclusion

This study highlights existing challenges in access to and uptake of vital postnatal health care within primary care, which were further exacerbated by the COVID-19 pandemic. Overall, fewer than half of women had a recorded postnatal examination within one year of delivery, with rates declining from 34.8% in 2013 to 22.2% at the height of the pandemic. Although attendance rates have partially recovered, significant inequalities persist across socioeconomic groups, ethnicities, and regions, with the most deprived and minority ethnic women consistently least likely to receive timely postnatal care. Addressing these barriers is essential to improving the quality and accessibility of postnatal care, particularly for vulnerable populations. Further research should examine the long-term impact of the pandemic on maternal health outcomes and evaluate the effectiveness of targeted interventions aimed at promoting equity in postnatal care and enhancing long-term health outcomes.

## Data Availability

No new data was generated by this study. The following existing data source was used: Greater Manchester Care Record available via https://gmwearebettertogether.com/gm-care-record/. The authors do not have permission to distribute the dataset generated by this study.

## Acknowledgements

This study was approved via the GMCR’s secondary uses and research governance process, which involved review against legal, ethical, and information governance criteria. The RECORD guidelines, a checklist devised for studies using routinely collected health data, were used to guide reporting.

The authors recognise the Greater Manchester Care Record (a partnership of Greater Manchester Health and Social Care Partnership, Health Innovation Manchester and Graphnet Health, on behalf of Greater Manchester localities) in the provision of data required to undertake this work.

---

**Supplementary Figure 1.**
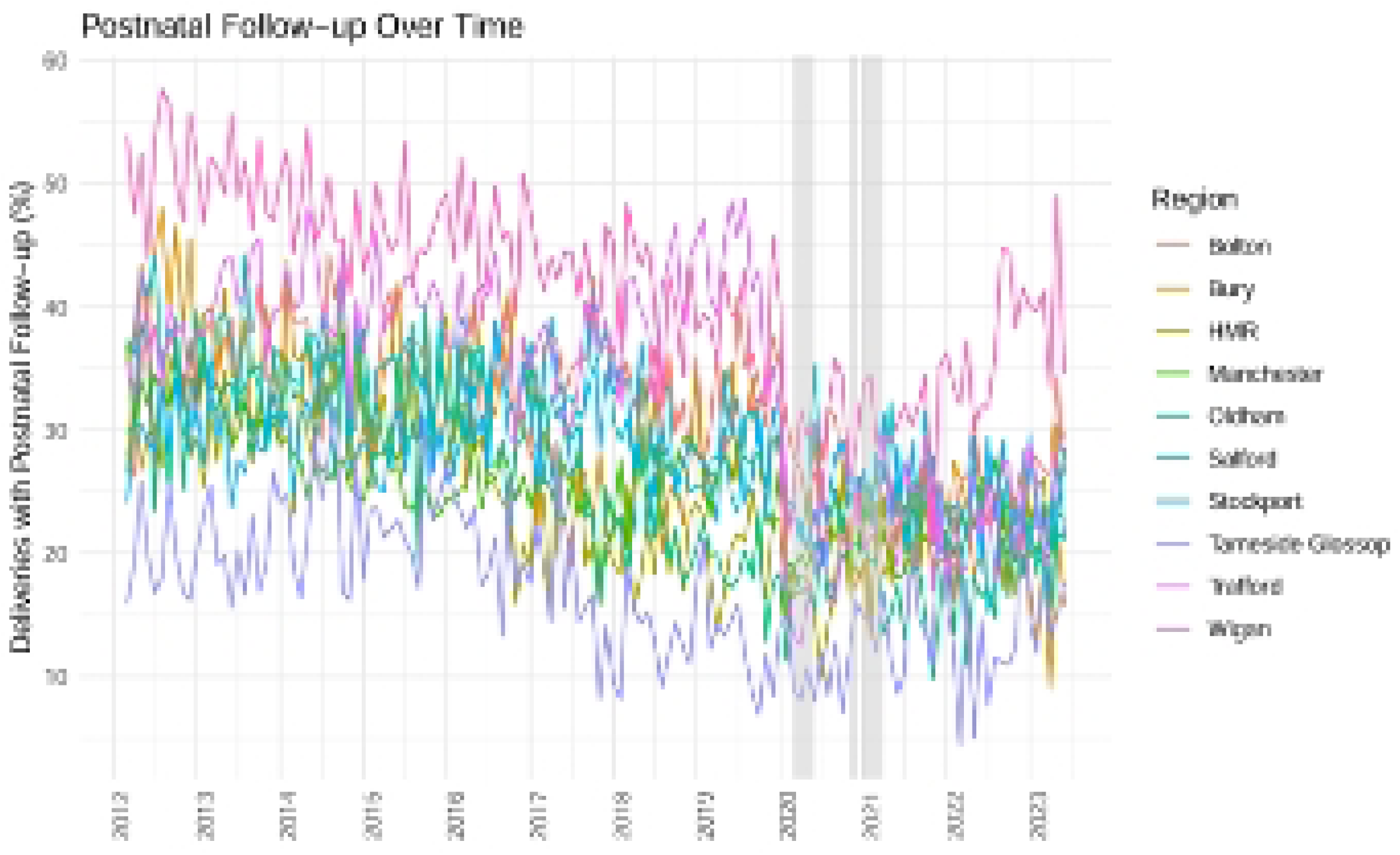

## References

1. Knight, M., Dealing with pregnancy problems—why we all need to be part of the solution. BMJ Medicine, 2022. 1(1).

2. Knight M, B.K., Tuffnell D, eds., Saving Lives, Improving Mothers’ Care - Lessons learned to inform maternity care from the UK and Ireland Confidential Enquiries into Maternal Deaths and Morbidity 2017-19. 2021.

3. NHS England. GP six to eight week maternal postnatal consultation – what good looks like guidance. 2023 [cited 2025; Available from: https://www.england.nhs.uk/long-read/gp-six-to-eight-week-maternal-postnatal-consultation-what-good-looks-like-guidance/.

4. National Institute for Health and Care Excellence (NICE). *Postnatal care NICE guideline: NG194*. 2021; Available from: https://www.nice.org.uk/guidance/ng194.

5. Department of Health and Social Care. Policy paper, Women’s Health Strategy for England (2019 to 2022). 2019 [cited 2025; Available from: https://www.gov.uk/government/publications/womens-health-strategy-for-england/womens-health-strategy-for-england.

6. Department of Health and Social Care. Women’s voices to be at the heart of renewed health strategy. 2026 [cited 2026; Available from: https://www.gov.uk/government/news/womens-voices-to-be-at-the-heart-of-renewed-health-strategy.

7. Yangmei Li, J.J.K., Christopher Gale, Dimitrios Siassakos, Claire Carson, Evidence of disparities in the provision of the maternal postpartum 6-week check in primary care in England, 2015– 2018: an observational study using the Clinical Practice Research Datalink (CPRD). BMJ Journal of Epidemiology & Community Health, 2021. 76(3).

8. NHS England. *Investment and Evolution:*. 2020; Available from: https://www.england.nhs.uk/publication/investment-and-evolution-update-to-the-gp-contract-agreement-20-21-23-24/.

9. Mesiano, N. and R. Santos, Impact of COVID-19 on primary care consultation mode in England: An interrupted time series analysis. Health Policy, 2026. 164: p. 105502.

10. Hayes, D.J.L., et al., The impact of the COVID-19 pandemic on the rate of maternal postnatal healthcare examinations in England: an OpenSAFELY interrupted time series analysis providing evidence of disparity in care access. BMC Med, 2025. 23(1): p. 626.

11. Manchester City Council, The Council and Democracy Census and Population, Census results 2021 | Census and population | Manchester City Council. . 2021: Internet.

12. Greater Manchester Sports Partnership, *Greater Manchester Socio-Economic Status Activity Levels Nov 17-18*, Greater Manchester Moving, Editor. 2018.

13. Health Innovation Manchester. *The GM Care Record*. 2020; Available from: https://gmwearebettertogether.com/.

14. Minassian, C.e.a., Methods to generate and validate a pregnancy register in the UK clinical practice research datalink primary care database. Pharmacoepidemiology and Drug Safety, 2019. 28(7): p. 923–933.

15. NHS England. *SNOMED CT derivative products*. 2023 [cited 2024 11/11/2024]; Available from: https://isd.digital.nhs.uk/trud/users/guest/filters/0/categories/8.

16. UoM-Data-Science-Platforms. UoM-Data-Science-Platforms, GM-SDE. 2022; Available from: https://github.com/UoM-Data-Science-Platforms/gm-sde/commits/master/projects/050%20-%20Palin

17. Institute for Government analysis. Timeline of UK government coronavirus lockdowns and measures, March 2020 to December 2021 2021; Available from: https://www.instituteforgovernment.org.uk/sites/default/files/timeline-coronavirus-lockdown-december-2021.pdf.

18. Brennan-Tovey, K., et al., Professional perspectives on barriers to accessing maternity care in England: a qualitative study. BMC Pregnancy Childbirth, 2026. 26(1).

19. Siobhán O’Connor, G.T., Olivia Jones, Anita Sharma, Laura Ormesher, Bradley Quinn, Anthony Wilson, Jenny Myers, Niels Peek & Victoria Palin, Acceptability of data linkage to identify women at risk of postnatal complication for the development of digital risk prediction tools and interventions to better optimise postnatal care, a qualitative descriptive study design. BMC Medicine, 2024. 22.

20. Claire X Zhang, M.A.Q., Clare Bankhead, Chun Hei Kwok, Nikesh Parekh and Claire Carson, Ethnic inequities in 6-8 week baby check coverage in England 2006– 2021: a cohort study using the Clinical Practice Research Datalink. British Journal of General Practice, 2024. 74.

